# Physicians’ learning in the workplace: use of informal feedback cues in daily practice

**DOI:** 10.1101/19010926

**Authors:** Carolin Sehlbach, Pim W Teunissen, Erik W Driessen, Sharon Mitchell, Gernot GU Rohde, Frank WJM Smeenk, Marjan JB Govaerts

## Abstract

**Purpose:** We expect physicians to be lifelong learners. Learning from clinical practice is an important potential source for that learning. To support physicians in this process, a better understanding of how they learn in clinical practice is necessary. This study investigates how physicians use informal feedback as learning cues to adjust their communication from interactions with patients in the outpatient setting.

**Methods:** To understand physicians’ use of informal feedback, we combined non-participant ethnographic observations with semi-structured interviews. We enrolled 10 respiratory physicians and observed 100 physician-patient interactions at one academic and one non-academic hospital in the Netherlands. Data collection and analysis were performed iteratively according to the principles of constructivist grounded theory. Our conceptual model describes how physicians use cues to reflect on and adjust their communication as well as to further develop their adaptive expertise.

**Results:** In addition to vast variations within and across patient encounters, we observed recurring patterns in physicians’ communications in reaction to specific learning cues. Physicians had learnt to recognise and use different cues to adjust their communication in patient encounters. They established a ‘communication repertoire’ based on multiple patient interactions, which many saw as learning opportunities, contributing to the development of adaptive expertise. Our findings show differences in physicians’ sensitivity to recognising learning opportunities in daily practice which was further influenced by contextual, personal and interpersonal factors. Whereas some reported to have little inclination to change, others used critical incidents to fine-tune their communication repertoire, while others constantly reshaped it, seeking learning opportunities in their daily work.

**Conclusions:** There is a large variation in how physicians use learning cues from daily practice. Learning from daily practice is a collaborative effort and requires a culture that promotes lifelong learning. Raising physicians’ awareness of experiences as potential learning opportunities might enhance their development of adaptive expertise.

## INTRODUCTION

Certified physicians are expected to engage in lifelong learning. As such, they are often required to prove their participation in formal learning activities, such as courses or congresses which too often are didactic and primarily knowledge-based (1-3). In fact, research findings suggest that most of physicians’ learning occurs informally through work (4-6). Compared to formal learning activities, informal learning is mostly unplanned, unconscious or tacit, and involves others through meaningful experiences in an authentic setting (7, 8). Informal learning can also be deliberate when physicians consciously aim to improve performance by, for example, seeking feedback or reflecting on experiences from their clinical practice, thereby engaging in self-regulated learning (4, 9-12). Feedback-seeking behaviour along with ongoing reflection on performance and performance feedback for the purpose of learning are at the heart of deliberate practice; fundamental to the development of professionals’ adaptive expertise (12).

Making informal learning more explicit and deliberate, therefore, remains imperative for physicians’ lifelong learning (4, 13). This specifically counts for further development and refinement of physicians’ communication skills. Communication is a complex yet crucial skill in patient care (14) that is predominantly developed through practice and informal learning.

Feedback and reflection on interactions with patients or peers in the workplace may stimulate physicians’ deliberate practice. It is well noted that certified physicians receive little formal feedback. Yet, informal feedback through other ‘learning cues’ including for example informal feedback from patient responses, clinical outcomes or conversations with colleagues, specifically reported by Watling and colleagues (15), may ‘‘facilitate the interpretation of the experience and the construction of knowledge from it’’ (15-17). However, it may be challenging for physicians to recognise those cues as cues for learning, as they can be ambiguous and are part of their daily routine (4). In day-to-day practice, physicians predominantly engage with patients using their routine expertise, while being less involved with their workplace learning (12). They may therefore benefit from support in how to recognise learning cues, as well as in knowing how to use and learn from them, so they may consider adapting and improving practice (2, 13, 15-17).

A better understanding of how physicians interpret meaningful experiences and use cues from daily practice as informal feedback may support strategies for physicians to develop their expertise, essential to ensure high-quality care (18). Our objectives, therefore, are to obtain an in-depth understanding of how physicians informally learn in and from the workplace (19). We aim to further refine theory on how physicians build and adapt expertise in the workplace setting (18). More specifically, we investigate how physicians recognise and reflect on learning cues related to their communication with patients and consequently, learn and adjust practice through patient interactions.

## METHODS

Using a constructivist grounded theory approach, we combined non-participant observations with semi-structured interviews (20).

### Setting and participants

We approached physicians from an academic and a non-academic hospital in the Netherlands. To ensure homogeneity with respect to the area of patient care, we selected respiratory specialists working with outpatients in respiratory clinics. Respiratory medicine includes a varied patient population including acute, chronic and terminally ill patients, which often results in long-standing and intensive physician-patient relationships. We purposefully sampled respiratory specialists with variation in age, gender, subspecialisation and experience. After the first round of data analysis, the research team decided to include additional physicians who were in the beginning, in the middle or at the end of their career for theoretical sampling. All physicians consented to participate.

Upon registration for outpatient appointments, patients received a short letter, which informed them about the research, the presence of a researcher as an observer (CS) and were asked for consent. The participating physicians verbally briefed each individual patient, emphasising that the research focused on the physician, that no patient data would be collected, and that CS did not have a medical background. If, after starting the consultation, either physician or patient preferred the researcher not to be present, CS left the room.

We obtained ethical approval from the Netherlands Association for Medical Education (NVMO: file number 2018.7.9), and from the ethical committees of both participating hospitals (file numbers 2018-0864 and nWMO-2018.118).

### Data collection

We combined data from non-participatory observations with informal and semi-structured interviews in an iterative design. CS shadowed 100 appointments of ten physicians in outpatient clinics. She observed physician-patient encounters and what cues physicians seemed to react to. She particularly focused on variations in communication styles during and across consultations and if, how and when physicians changed their communication with patients. During the observations, CS took field notes using ethnographic techniques, which she worked out after the observation within the following 48 hours. Observations lasted 1.5 to 3.5 hours, during which CS usually observed eight to twelve outpatient appointments (Table 1). CS sat with some distance from the physicians and patients, and asked participating physicians to conduct the clinic following a typical daily routine.

**Table 1:** Overview of physician-patient interactions observed

| Interviewee | Number of Patient encounters | Number of hours observed |
| --- | --- | --- |
| A1 | 12 | 3 |
| A2 | 8 | 2,5 |
| A3 | 11 | 3,5 |
| A4 | 11 | 2,5 |
| A5 | 12 | 3 |
| A6 | 11 | 3 |
| A7 | 11 | 3 |
| A8 | 10 | 3 |
| A9 | 10 | 3 |
| A10 | 4* | 1,5 |
| **Total** | **100** | **28** |

\*One observation included four patient encounters in a highly specialised outpatient clinic for patients with a certain rare disease.

After the observations, CS used her notes to prepare interview questions on situations observed. Following this preparation (0.5 to 1.5 hours), CS conducted semi-structured interviews. During the interviews, physicians considered and reflected on cues they used to adjust their communication. First, CS inquired about changes she had observed in physicians’ non-verbal or verbal communication during specific patient encounters. Second, her questions addressed differences in how physicians approached different patients. Drawing on these observations, CS probed the physicians interviewed on their awareness of the learning cues that led them to alter communication during interactions with patients. Semi-structured interviews lasted 30 to 55 minutes, and were recorded and transcribed verbatim. With the help of the transcriptions, notes were transformed into concrete reconstructions with analytic memos and commentaries (21). The data were subsequently analysed to obtain an in-depth understanding of the cues physicians considered relevant to their communication, and how they used these cues for learning.

### Data analysis

Data collection and data analysis were performed in an iterative manner, so that early analysis influenced the focus of the following observations and interviews. CS started by open, line-by-line coding of three interviews and the corresponding observational field notes. The authors FS, ED, PT, GR and MG each read and coded one interview transcript and related field notes and discussed their respective codes with CS independently. After refining the coding framework, the research team discussed and collated codes into preliminary categories. PT and CS jointly discussed these, which resulted in the following preliminary categories: context, physician-patient relationship, patient characteristics, anticipating or reactive change in communication, and routine. CS used the preliminary categories in focused coding of two more interview transcripts, before discussing categories with SM and PT separately. This led to the construction of the following additional categories: external factors, examples of learning cues (patient reaction, physician reaction), reflection, and communication repertoire. CS and SM conceptualised the categories into a conceptual model, based on which CS coded three more interviews. CS, SM, FS, ED and MG discussed and agreed on the conceptual model, and further deliberated how contextual factors affected physicians’ decisions to react to and learn from cues or not. After CS had coded the two remaining transcripts, the research team re-examined the conceptual model and adjusted it into a final conceptual model of how physicians recognised and used informal feedback cues for their learning and how using cues resulted in a change of communicative behaviour.

We collected and analysed data until the authors felt that no new concepts came up and theoretical sufficiency was reached to answer the research question and to build theory (22, 23). Data was managed with ATLAS.ti (ATLAS.ti Scientific Software Development GmbH, Berlin, Germany) and the analysis reported with the COREQ checklist (Appendix I).

### Reflexivity

The research team maintained reflexivity throughout data collection, data analysis, and writing up the results by discussing underlying assumptions about physician-patient communication and learning cues. CS used reflective memos throughout data collection to reflect on how her own experiences and pre-constructed knowledge influenced the interpretation of findings. She has a background in health sciences and is a PhD student in medical education, focusing on physicians’ lifelong learning. The research team also consisted of medical educators (MG and ED) and medical specialists (FS, GR, PT). FS and GR are experienced respiratory specialists. PT is specialised in gynaecology and obstetrics. SM is a medical education manager. All members of the research team are involved in medical education, having a special focus on workplace learning, continuing education or performance assessment. Their respective backgrounds might have influenced the research in presuming that physicians learn from daily patient encounters.

## RESULTS

Our analysis revealed differences in how physicians reacted to, reflected on and learnt from various cues related to outpatient communication. The table below underlines potential cues that participants identified and the researcher observed (Table 2).

**Table 2:**
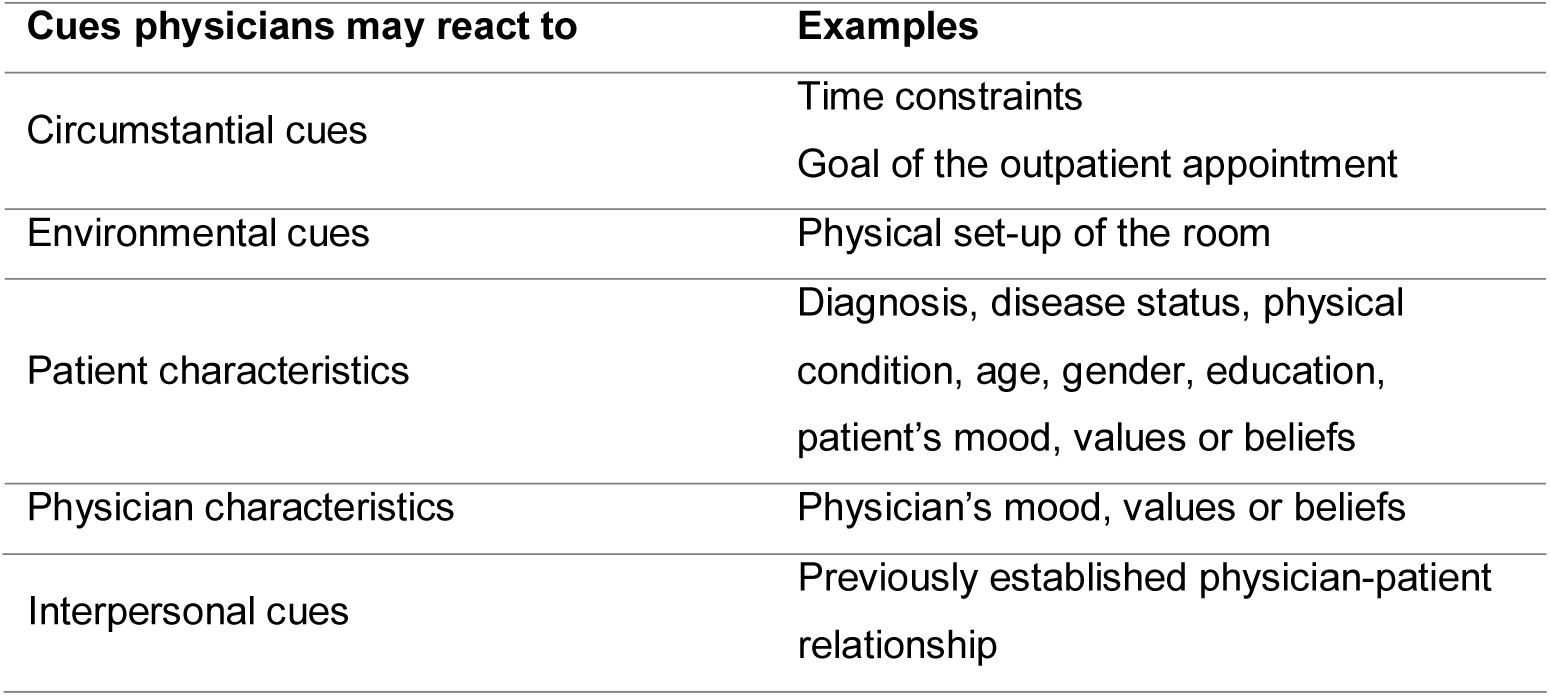
Examples of different cues in patient interactions

In the following paragraphs, we first describe the variation in the physicians’ communicative behaviour we observed within and across outpatient appointments. Subsequently, we report how physicians felt they learnt or did not learn from opportunities in their current practice.

### Variation in communication within and between patient encounters

Physicians faced large variations of interactions in the outpatient setting. We noticed recurring behaviours in physicians’ communication, which they indicated to have acquired from previous interactions.

Physicians’ communication differed between patient encounters. They continuously adjusted to patients’ reactions and circumstantial cues, smoothly manoeuvring through different patient encounters. One physician portrayed this as ‘the game in the consulting room’ (observational field notes, A7). They had a ‘communication repertoire’ at their disposal, which guided them through their daily routine while either deliberately or unconsciously reacting to context-specific cues:

> It is actually just one strategy: … that is, responding to what the patient gives you back. That can be really easy sometimes and passes very quickly, but other times you really need to manoeuvre carefully I now have a huge repertoire of standard reactions that I can draw on …. (Interview, A4)

Physicians’ communication repertoire included, but was not limited to, different strategies of taking the lead in the conversation, drawing to illustrate or explain a diagnosis, prognosis or treatment, change in body language and/or non-verbal communication. Some observations showed, for instance, that physicians leaned forward to show interest in and empathy for the patient or to emphasise the severity of a patient’s condition. Some used physical contact to comfort or console patients by shortly touching an emotional patient’s hand or patting them on their back when saying goodbye. Similarly, physicians signalled the end of a consultation by leaning backwards, pushing back their chair or standing up.

Our participants realised that different factors, such as the atmosphere or goal of the consultation, the physical space of the clinic and time pressure, added complexity to the outpatient appointments. Recognising these circumstantial cues, they understood that patients might be agitated when coming for a test result or check-up after a treatment and had learnt to adjust their communication accordingly. One interviewee, for example, gauged the atmosphere during a patient encounter for deciding, deliberately, whether to open or wrap up a consult or reassure patients with a joke: [The physician jokes:] ‘With this, you should be able to live to be a 100. But I cannot give you any guarantee, of course.’ (Observational field notes, A4) The same physician had made identical, age-related jokes in previous consultations. In another consultation, however, he remained distant and refrained from making jokes, because, as he explained afterwards, ‘this was not the most cheerful patient’. These observations underline that, depending on the circumstances, physicians deliberately adapted their communication. Along with circumstantial cues such as atmosphere, workload or time constraints, interpersonal and personal cues also affected physician-patient communication, including the relationship previously established, physicians’ and patients’ mood as well as their norms, values and attitudes:

> You only really get to know people if you see them often. … At a regular check-up - I do think that when you see familiar people … that I pay more attention to ‘What kind of person was that again?’ … That is, I think, your mental preparation, and I think at that point you are already largely determining your communication strategy. (Interview, A2)

We observed how physicians adjusted their communication while anticipating their patients’ reaction by slowing down, leaving a silence, increasingly using gestures or giving more detailed information. We could distinguish between *ad hoc* adjustments in reaction to an unexpected turn of events such as a patient bursting out in tears or a phone ringing, and planned behaviour in anticipation of the consultation flow (i.e. breaking bad news), as in the observed consultation described below:

> A cancer patient and his partner return to discuss treatment options, because their son had expressed his dissatisfaction about the diagnosis and treatment option the trainee had given his father. The trainee asked the attending physician to lead the consultation and to discuss the diagnosis and treatment plan with the patient and his partner.

> The physician explains the diagnosis. He speaks calmer than before when listing treatment options, pauses and gesticulates with his hands. ‘Let me draw it. You will receive more information about this later, but schematically for now…’

> The phone rings twice during the consultation. The physician silences it both times, the second time even without looking at the phone or pausing his explanations. When there is a third call, the trainee takes it.

> The phone also rang during other consultations that day, but this time the physician did interrupt the ongoing conversation to answer it.

> (Observational field notes, A7)

Physicians had developed a communication repertoire that enabled them to quickly adapt their behaviours depending on how various interrelated factors influenced the patient consultation.

### Learning from daily practice

Our analysis of the observation and interview data revealed two possibilities of how physicians applied their communication repertoire in daily practice, recognising learning opportunities in past or present patient encounters. Although all physicians had acquired a daily routine, some were able to uncover learning opportunities within this routine and engaged in ongoing expertise development.

Some physicians felt that they were no longer able or willing to learn or adjust. They felt that time constraints and daily routine took priority over using patient encounters as learning opportunities to adapt their communication. They remarked to be ‘stuck in this pattern’ (interview, A5), preventing them from learning communicative subtleties, and described that the ever-repeating routine had decreased sensitivity and willingness to learn from their interactions with patients. Reflecting retrospectively on their communication helped them realise how they had acquired their behaviour. Our interviews forced physicians to deliberate on the preceding patient encounters and potential learning opportunities. They were unconscious of their communication repertoire and how they used similar strategies across patients: ‘How funny. … It could be. … I didn’t know that I pushed my chair back, but now that you mention it, I think, oh yeah, that’s right, I do that.’ (Interview, A8).

Interviewees repeatedly mentioned that an absence of patient complaints implied that their communication repertoire was sufficient with no need for further development. They heavily relied on the repertoire built and believed they could ‘no longer be corrected’ (Interview, A5). While comprehending its importance, physicians hardly dwelt on their communication during or after patient encounters. Some struggled to pinpoint how they adapted their communication throughout patient encounters, which reflects an automatic behavioural adjustment. Others knew they used humour in their consultation but often mentioned it as an unintentional and automated strategy:

> That way you try to keep it a bit personal, to put people at ease perhaps. … But that is something that you sense unconsciously. It’s not as if I think like ‘oh, let’s make a joke now’, it’s something that happens unknowingly, actually (interview, A10).

Even stronger, many considered their communication repertoire as part of their personality: ‘That’s how I am. Some [physicians] are more cheerful or more business-like. It’s not something you can switch on or off’. (Interview, A1). As one of our respondents explained:

> If this is the right way or [if] it could be better, yes, that may be so, but this is *my* way. And I don’t think that I’m doing it a lot differently now from the way I did it 10, 20 or 30 years ago. (Interview, A5)

We found a notable contradiction in this response as the same physician claimed to still learn:

> You know, frankly, that sounds very arrogant, but I don’t think I’m doing a very bad job. But I know for sure that sometimes I can be entirely wrong. And I do try to learn from that. It touches me. It certainly touches me. (Interview, A5).

For this physician and most of his colleagues, ongoing learning does not represent a smooth learning curve, but occurs intermittently induced by emotion or critical events. Some indicated to learn from difficult or peculiar patient encounters, which were out the scope of our observations. They reported that those learning events often stood out from daily routine, ‘ranging from someone who [is holding] a chair above his head and threatening you, so to speak, to people who … well, all sorts of variations’ (interview, A7). Critical incidents resulted in increased reflection which made physicians question their own behaviour and re-evaluate their communication repertoire. One of our physicians painfully described his communication as a shortcoming, because his patient continued smoking after repeated warnings. He believed to ‘be failing as a doctor’ (interview and observational field note, A6) and recognised that he needed to learn how to deal with patients who trivialise their condition. This shows that despite having developed a reliable repertoire, physicians’ learning behaviours may change when experiences elicit emotions, especially when they feel to have failed.

Other participants internalised regular reflection as part of practising, continuing improvement and learning from their practice. These physicians readily acknowledged that their communication repertoire was ‘based on previous experiences’ (interview, A4). They were aware of the constant yet subtle adaptations in their behaviour within and between patient encounters. Physicians used these to play ‘the game in the consulting room’ of deliberately fine-tuning and mastering their communication repertoire: ‘You try to act on the patient’s level. This morning, we had that farmer, right, that man who reacts primarily driven by his emotions with a certain rigidity and then you try to tune into that level’ (Interview, A7). Some indicated that reflection on and learning from patient encounters had become fully ingrained in their daily practice: ‘I’m always, I think, learning a bit anyway’ (interview, A8). As someone explained in reaction to our observations:

> Yes, I do think that I actually always take note, when the patient [is] in the consulting room or [when I] start preparing for the next patient. Like, … for this patient, but also for similar cases, … how do I say that in plain and clear language? … I did learn from this and reflected [on it] again and that’s actually how I do it every time. (Interview, A2)

## DISCUSSION

We aimed to explore how physicians use cues from their clinical practice as informal feedback to adjust their communication and learn from daily patient interactions. Our data showed variation in physicians’ communication patterns within and across outpatient encounters influenced by personal, interpersonal and contextual factors. Physicians reported to have learnt from previous interactions, which established a communication repertoire that guided them through their practice. We described different degrees of the extent to which physicians recognised and used informal feedback from daily practice as cues for learning and learning opportunities.

The two opposites we present show that some physicians are less likely to engage in reflection and deliberate learning from day-to-day patient care, whereas others appeared more sensitive to learning cues to reflect and adapt their communication. Some of our participants were caught up in the demands of daily practice and did not have the time or willingness to reflect on their communication repertoire. According to Ericsson’s model of deliberate practice and expert performance (9), these physicians were in a phase of arrested development. Yet, they felt competent in their performance and would potentially reinitiate a learning process in response to critical events. Others continuously challenged their communication repertoire to ultimately increase control over unknown situations (24). They reflected on whether they had used their repertoire effectively and efficiently which allowed them to smoothly navigate through practice as an indicator of mastery and expert performer (9). This variation in physicians’ willingness to lifelong learning reflects on previous findings on expertise development (9, 25). Mylopoulos et al. (25) argue that adaptive experts may not only recognise the “old in the new” but that they also reconceptualise their practice by reflecting on the “new in the old”. This echoes our findings that some physicians may regard patient encounters as learning opportunities to confirm or fine-tune their communication repertoire. It also induces the questions about which factors influence physicians’ position on the learning curve, or how we can create a stimulating work environment for all physicians to become lifelong learners.

The way participants in our study described and reflected on the development and use of their communication repertoire reflects findings from a recent study on how trainees develop expertise in communication (26). Kawamura, Harris (26)and colleagues (26) distinguish between procedural fluency and conceptual understanding in expertise development in communication. The use of humour or physical contact to comfort a patient can be considered procedural fluency, whereas ‘shifting’ between patients and moving towards patients’ ‘level’ reflects a physician’s conceptual understanding of distinct patient needs (26). Adaptive experts thus command a conceptual understanding of when to use their established communication repertoire and when to adapt how they communicate their clinical knowledge to different patients or when to innovate strategies for adapting to non-routine situations. Findings from our study seem to confirm Kawamura and colleagues’ conclusion that learning how to ‘shift’ between patients is key to expertise development in communication.

The degree of willingness and sensitivity to recognise learning opportunities that we describe, stress Teunissen and Bok’s reflection on goal orientation and self-theories (27). Their distinction between incremental and entity theorists shows parallels to our results (27, 28). Physicians who reflected on their actions and emotions, and who continuously tested their communication repertoire showed signs of a growth mind-set. They appeared as intrinsically motived lifelong learners and adaptive experts. Among our participants, we, however, also recognised individuals who were more performance-oriented. They were mainly absorbed in the acquired routine and considered the absence of patient complaints as affirmation of good practice. Possibly, contextual factors within daily practice had diminished their incremental mind-set over time which hints at possibilities for improving the overall learning climate in hospital departments (28). Engaging physicians in lifelong learning may require a culture in which the tension between performance and continuing learning is recognised. When physicians are given the opportunity to reflect more on their communicative behaviour during patient care, through formal and informal feedback, a growth-mind-set may be stimulated.

Contextual factors such as workload and time pressure in the clinical workplace may impede physicians’ reflection on learning cues and their learning (17, 29-31). Some of our participants were more sensitive to time pressure and preoccupied with daily routine than others, which may

explain the individual differences in how deliberately our participants recognised learning opportunities. This aligns with the findings of Kyndt and colleagues (30) who suggest that learners’ characteristics as well as organisation type and size determine the acknowledgement of learning opportunities at work. Engaging physicians in lifelong learning may thus require a culture which pro-actively stimulates reflection and the development of a growth mind-set and adaptive expertise.

### Implications in relation to lifelong learning

Our results indicate that we may need to support physicians in engaging in reflection on action, while being cautious of potentially negative effects. We need to balance routine practice with reflective practice (to support development and maintenance of expert performance) since continuous reflection may hinder physicians in delivering care (9). That is, physicians need to have a certain level of automaticity to practise considering the increasing practice demands and contextual factors such as workload and time pressure.

Nonetheless, if feedback and reflection are truly valued as essential attributes for lifelong learning, more importance must be placed upon physicians’ awareness of their own behaviour during practice, combined with a willingness to learn in order to improve. The UK revalidation system, as one of few national recertification systems, already requires physicians to reflect on critical incidents, feedback received, and complements or complaints during an annual appraisal. Although this creates workplace-based opportunities for physicians’ learning, physicians are likely to reflect on aggregated data only, de-emphasizing learning-in-context. Rather, if we want to stimulate more reflective behaviour in practice, we may consider it as a collective effort by creating an environment that stimulates learning. Physicians could, for instance, occasionally observe peers to face diverse communication approaches to learn for their own practice, similar to how trainees learn from observing faculty (26, 32). To initiate collaborative learning, physicians could also engage in peer consultation and share experiences and recent learning as described by Mylopoulos et al. as “discover then tell’ (25). Those who are capable of discovering learning opportunities among patient interactions, could guide their colleagues who are less sensitive to cues in a collective effort to engage in deliberate practice (9). Perhaps some of these collective learning activities could then serve as evidence that a physician contributes to a learning environment as a marker for recertification.

### Strengths and limitations

A first limitation is that we base our data on single day observations and self-perceived learning strategies. We did not observe the same physician for a longer period of time. Following a number of physicians longer, would have resulted in multiple observations on physicians’ learning behaviour, also outside of the outpatient setting. We collected data in outpatient clinics for respiratory disease, observing and interviewing respiratory specialists. Both hospitals were teaching hospitals. This might present another limitation as physicians working there might be more aware of and involved in others’ or their own learning due to their teaching role. These physicians might have (developed) a more learning-focused mind-set than physicians without educational duties. It would be worthwhile exploring how physicians learn informally in a non-teaching context. In the Netherlands, recertification procedures and requirements to guide physicians’ lifelong learning are similar across medical specialties. This makes our results transferable to other specialties, particularly considering that variations between and within individuals are presumably present across all specialties. Research in other specialties or clinical contexts and settings, however, may lead to different findings.

A strength of our study was the combination between non-participant observations and interviews. Many others have previously recommended this combination of methods to present data on what physicians report themselves but also on what can be observed (11, 17, 18). Observing physicians in the outpatient setting was instrumental to help us to explore learnt behaviour and learning opportunities in practice.

## Conclusion

There is a large variation in how physicians use learning opportunities in outpatient settings. Their informal learning is influenced by contextual, personal and interpersonal factors, which might either promote or inhibit physicians’ reflection and learning. Our findings suggest that there is some importance that can be attributed to making physicians more aware of when and how they can learn from daily practice, also in collaboration with others. Learning from daily practice is a collaborative effort and requires a learning culture, in which physicians can use existing differences between their own and their peers’ performance to learn from each other.

## Data Availability

The data that support the findings of this study are available on request from the corresponding author.

## Notes

### Competing Interest Statement

The authors have declared no competing interest.

### Funding Statement

The European Respiratory Society funded this study. However, Carolin Sehlbach, who was appointed to the research project as a PhD student at Maastricht University conducted the research independently of this funding. The European Respiratory Society had no role in the design or conduct of the study; in the collection, analysis, and interpretation of the data; or in the preparation or approval of the manuscript or the decision to submit for publication.

